# Women Empowerment in Medical Imaging and Radiology: Status and Challenges

**DOI:** 10.1101/2024.01.03.24300758

**Authors:** Ashwag Rafea S Alruwaili, Alanoud Abdullah Alzahrani, Hajar Abdullah Almater, Lina Abdulaziz Aldibas, Nawal Farah Al anazi, Asma Mohammed Albanyan, Wadha Alyami

## Abstract

Since the pandemic’s beginning, there have been significant setbacks in social and economic inclusion of women. The resistance by public can be quite challenging too due to different reasons, such as religious beliefs or the dominance of custom and traditions creating a consistency in leadership perceptions and prototypes of leaders. The Saudi vision unleashed the exacerbated efforts to increase women involvement in the workforce and empower them in leadership roles. Yet the overall representation of women in leadership positions in health and academe in Saudi Arabia remains low compared to other countries. Exploring women status in medical imaging and the risks of reversals after the pandemic became necessary. An online questionnaire was distributed to female workers in both health and academic institutions in the field of medical imaging from December 31, 2021 to October 10, 2022. A total of 250 responses were received, and only 240 females consented and met the criteria for inclusion in the study. Results show that Saudi women still face the glass ceiling when it comes to administrative roles even though they have high level of self-esteem and efficacy along with the support from the Saudi government by fair wages compared to male counterparts. However, institutions imposed largely more constraints on women, creating difficult environment for gender diversity in work culture such as breastfeeding hour, rest during working hours, leave for accompanying family member in sickness or hospitalization such as children, sick leaves during pregnancy or monthly period. In a remarkably short span of time, the current Saudi government has undertaken commendable initiatives to empower women, achieving their predetermined targets with notable success. Despite that, there are many familial and cultural challenges that hinder them from leading positions. The perceived efficacy of women leaders and the disparities in their leadership philosophies are still up for research.

## Introduction

Global development for women and girls has halted due to the economic and social effects of the pandemic and geopolitical strife, which also run the risk of leaving permanent scars on the universal movements towards empowering women. Empowerment is a combination of power and self-sufficient rather than a single process only, that stimulate individuals to positively contribute to the success of communities. Qualified females remain marginalized in leadership and administrative positions [1]. According to the United Nations Educational, Scientific and Cultural Organization (UNESCO), the global science records showed less than 30% of researchers are female, in which there has been simply 42% in Asia and 30% in Africa, and 32% in North America and western Europe, and 45% in Latin America [2]. This may explain why female deliver out much less scientific papers than male [3]. In the United States of America (USA), the presence of female in decanal positions (ie, dean, vice dean, senior associate dean, associate dean, assistant dean) in medical schools make up at most only 15% of the deanship population [4]. In Saudi Arabia, female researchers represented even less than the numbers by UNESCO where only 23.2 % of total researchers are female [5]. The gender prejudices and constrictive gender norms that exist in society are reflected and reinforced by the health systems too [6]. This evidence of bias endangers the well-being of female healthcare professionals as well as the health of entire communities.

Culture can be one of the barriers to women empowerment. The belief of the community that health workers should be driven by altruism rather than financial gain is encouraged by health professional themselves and culture norms. This belief can justify the low-paying or unpaid roles, such as volunteering, for females in the health sector. Some collectivist culture, such as middle eastern, public take enormous pleasure in upholding and preserving the Islamic and Arabic traditions and might not be prepared for the change towards women leadership. Particularly, in the Saudi culture, Saudi citizens nurture the Islamic root of their culture which mistakenly misbelieve that women cannot lead [7]. Although since the announcement of Saudi 2030 vision, women were the highlight of the media focus. Yet empowering women face many obstacles and this quantum leap in new social trend might reshape the Saudi culture in the coming years. Further challenges such as environmental factors, familial responsibilities and duties, male worker dominance at workplace, and gender equality [8]. The lack of self-motivation and level of education are found to be more personal challenges [9]. Extreme issues include sexual harassment in workplaces [10], lack of voice, image of incompetence and male co-workers discomfort with women whom they perceive to be better than them [11] were previously reported and recognized. There is evidence that entry of women into the field of medicine is about 50% [12] while the role of women in leadership is way less due to reasons such as delayed leadership opportunities, gender inequality and unequal capabilities. Some female faculty members may have difficulty combining professional and family responsibilities, leading them to leave their leadership work in favor of male due to a lack of cooperation and work-life balance [13]. Empowering women is derived from the societal concept that this gives women their necessary rights in life, equally to education, making their own decisions, and not being subject to the dominance of masculinity [14, 15].

It has been found that even with rising reports of women experiencing violence and discrimination [16, 17], empowering women will always provide safety and protection for female to resist the abuse that goes toward them [18]. Fundamental problems can be addressed through awareness and change in institutional policies towards the role of women, equal opportunities, cooperation to resolve work-and-life imbalance and therefore, improvement in the development of leadership skills.

There have been recent efforts to increase women involvement in the workforce and empower them in leadership roles [19, 20]. The overall representation of women in leadership positions in Saudi Arabia remains low compared to other countries. Based on the Global Gender Gap Index, Saudi Arabia held the place of number 141 out of 149 in 2018 [21]. Despite the 130% increase in the number of women working in the private sector by 2022, their leading participation still lags behind their political empowerment and education indices (127 out of 149) [21]. Further research is needed to fully understand the challenges and obstacles hinder the progress of women empowerment by 2030.

There is a scientific agreement on the biological differences between women and men which cannot be neglected. However, the cultural differences are manageable and can be alleviated. Achieving gender equality in Science, Technology, Engineering and Medicine (STEM), can bring us to the ideal health, communal, and economic growth. Gender equality help not only in economic expansion, but lowers the death rate in children, and helps build up nutrition [22]. Worldwide, repression of women has serious consequences on physical and mental health [23].

The historical stringent restrictions on Saudi women such as mobility freedom compared to Gulf region was not restricting Saudi women from taking the lead among the gulf women in directing the system’s performance towards health and education with the government’s support to meet women’s rights and needs [24]. The changing in the policies of Saudi Arabia is in parallel to the community and society increased awareness towards women’s education and rights which are important elements that will improve women’s self-efficacy [25]. The new vision of Saudi Arabia promotes women empowerment through relaxing the constraints placed against them. It considers women a great asset and has been publicly published that female’s presence should take part in 30% of leading presence in Saudi system since 2016 [7]. Since the launch of the Kingdom’s Vision 2030, women participation rate increased from 21% in 2017 to reach 31.2% of the workforce in 2020 [26]. The reforming of the whole system, whether its education or health systems, are in favor for women to play a real role in the advancement of the lifestyle in the Kingdom. These recent moves hold important lessons to put more effort on emerging fields such as medical imaging in both medicine and academe [27]. Establishing new policies in order to ease women’s career life are very crucial at this stage of transition towards women empowerment. Especially with the aim of lifting restrictions on women to join sectors that previously considered male specialty and to enabled women to access leadership positions. Therefore, the need to investigate women status in the health and academe of Radiology field is demanding. The changes implemented to women empowerment in medical field over the past years remains unprecedented. However, there is still uncertainty about the direction of leadership opportunities and the full empowerment of women in the fields of Radiological sciences and medical imaging. This study aimed; a) Investigate women’s status in leadership and administrative positions in medical imaging field from research the female current status in medical imaging, b) explore other empowering factors related to women’s leadership and administrative roles, c) to address the difficulties facing female empowerment in medical imaging, d) to provide recommendations that can aid the health system towards women empowers in medical imaging.

## Methods

### Sample size

The recruitment period commenced on December 31, 2021 and concluded on October 10, 2022. During this timeframe, participants were actively sought and enlisted for inclusion in the research, ensuring a comprehensive and representative sample for the investigation. This survey-based study received responses from a total number of 250 and included only 240 female who consented and met the criteria for inclusion; a) Saudi female, b) aged 25 years old or above, c) working in the medical imaging departments or radiological sciences departments either in hospitals or universities in Saudi Arabia.

Participants should hold an education degree either a Diploma, Bachelor’s, Master or Ph.D. workers and academics in the field of medical imaging. Results were based on their perspective towards women’s empowerment and the difficulties facing them against their empowerment and what is required to empower them.

### Questionnaire design

The bilingual online surveys were designed by researchers in English and Arabic and divided into four sections: a) demographic data which will cover basic information about the region, age, marital status, number of kids, education level, sector, size of the hospital, years of experience, monthly income if they held a leadership position, and the current position, b) personal empowerment which includes questions about self-esteem, and self-efficacy, c) social/ relational empowerment which includes statements about economic empowerment, and freedom of mobility, d) environmental/ workplace empowerment access to education, resources, support, organizational role, and women rights in work.. A copy of the questionnaire is available in supp 1.

The questionnaire was designed to include several domains that are classified under two dimensions, as follows; The **first section**: include multiple domains with factors related to leadership and administrative roles for women in the field of medical imaging which is divided into several dimensions. The **second section**: difficulties that face females in the medical imaging field. The next step was formulating the statements of the questionnaire. After reviewing the past theoretical models, research and previous literature related to woman leadership in medical imaging, with the tools used in previous studies, we constructed current tool statements. A 5-scale Likert scale has been set for responses where one is strongly disagreed and 5 represent totally agree.

### Statistical analysis

Exploratory data analysis and cross tabulation was carried out such as calculated frequencies of participants for each question, weighted average, and standard deviation. Both percentage and frequencies were applied to identify the demographics of the study sample and to determine the results of the main dimensions in the study tool. Weighted Mean was used to investigate the level of agreement on the statements and arrange the statements based on the highest weighted mean. Validity of the study tool and internal consistency are available on Supp 2

### Ethics

Ethical approval was issued by the Institutional Review Board (IRB) at King Saud University Medical City (KSU-MC) number E-21-6532. Participants provided informed consent by answering an explicit question before proceeding with the survey, affirming their voluntary participation and understanding of the study’s objectives and their rights.

## Results

### Study population

The sample represents the female community of all specializations in the field of radiology in both health and academic sectors. The questionnaire was distributed online to the study population, and a total number of 240 participants were valid for statistical analysis.

### Sample demographics

Geographical Distribution-Most of the respondents are from Riyadh represented by 55.4% of the total study sample, and they are the largest category of the study sample. The next dominant percent was represented by the Eastern region of 13.8 % while the lowest percentage of participants was from both Albaha (0.8%) and the Northern borders (0.4%). The regions of Asir and Makkah had almost similar numbers of respondents (7-7.5%). Figure 1 shows the geographical distribution of the study sample and the years of experience. Most of the respondents had 5 years of experience or less (n=145, 60.4%), and they are the largest category of the study sample. Next, is 6 - 10 years of experience (n=52, 21.7%), then the 11 - 15 years of experience (n= 23, 9.6%). Only 20 participants had over than 15 years of years of experience, representing 8.3 % of the total study sample.

**Figure1.**
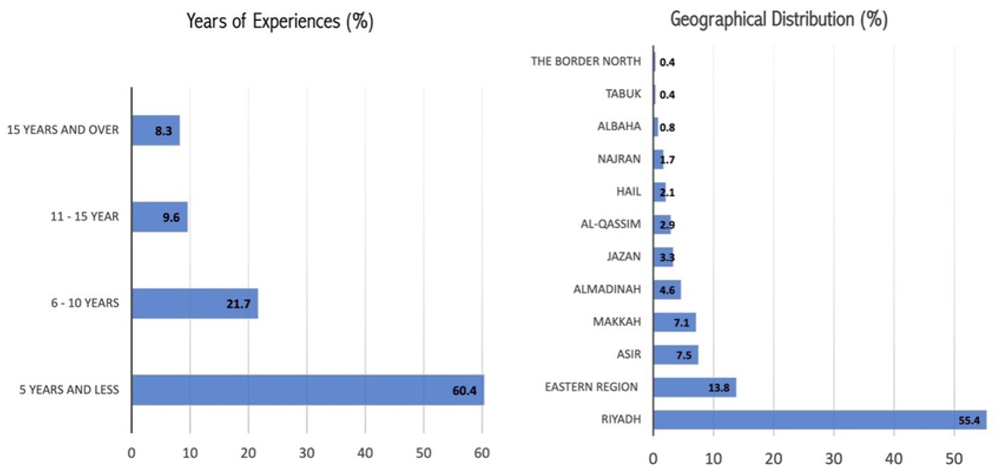

Age groups-the study sample’s age group of less than of 25 years old (n= 118) represent 49.2% of the total study sample’s age group and they are the largest category of the study sample. The respondents who are between 25-30 years old (n= 44) represent 18.3% of the total study sample. The group age of more than 30 to 35 years (n= 43) represents 17.9% of the whole respondents. Above the age 35(n= 35) represents 14.6%. Table 1 shows the details of each age group.

**Table 1:**
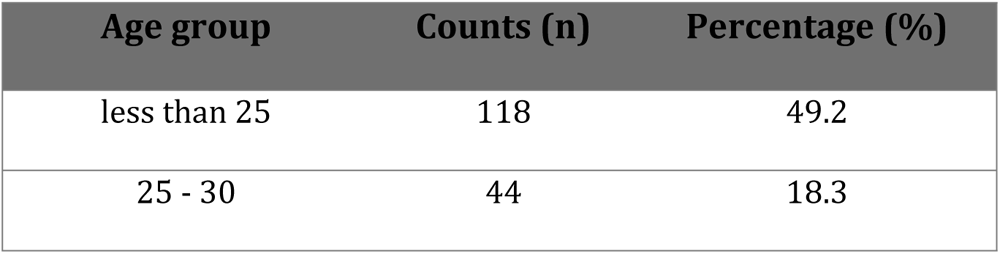
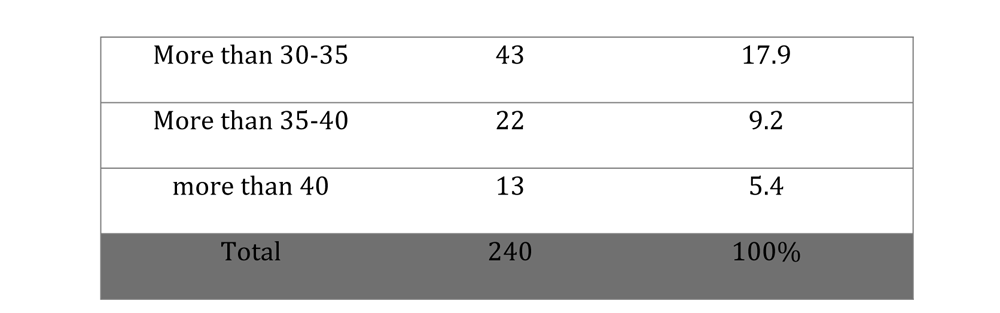
Age distribution of the total participants.

The majority of the study sample’s (n=125) are single females who represent 52.1% of the total respondents and have never been married. While (n=108) of them are married and represent 45% of the female sample. The divorced females (n=7) are the lowest number of the study sample representing 2.9% of the total number of the study sample. Among the last two samples of married (n=108) and divorced (n=7), A total of 70.4% of (n=81) have 1-2 children, 28.7 % (n=33) had 3-6 children, and only one participant had more than 7 children. The annual income of most respondents (n=176, 73.3%) is equal or more than $32,000 and they are the largest category of the study sample, while only (n=38,15.8%) of them represent the sample who had their annual income of $16,000 to $32,000. While the least are the female respondents (n= 26, 10.8%) who receive an annual income of $16,000 or less, (Figure 2-A). The sample education level distribution was a follow; respondents with undergraduate degree (n=181, 75.4%), master’s degree (n=26, 10.8%), diploma (n=23, 9.6%), and Ph.D. (n=6, 2.5%). Only (n=4, 1.7%) were in fellowship (Figure 2-B) shows the education level distribution for the sample included in this study. More than half of the respondent (n=160, 66.7%) are representative of the government health sector of the study sample and (n=39, 16.3%) in the private health sector. Among these groups, (n=44,22.1%) worked in a hospital sized 100 beds, (n=29, 14.6%) in hospitals sized 110-150 beds, and majority of the health sector respondent (n=126, 63.3%) worked in bigger size hospital of more than 150 beds compared to the others (Figure 2-C). Figure 2-D shows the employment field and the size of hospitals for the respondents from health fields. This study found that only (n=44, 18.3%) of females have been in leading roles or administrative positions while the majority (n=196, 81.7%) have never worked in a leadership or administrative position. From the 18.3% female leaders, majority (n=32, 72.7%) held the leadership/administrative role within the medical imaging field and only (n=8, 18.2%) of them, held these positions outside the medical imaging field. Only few (n=4, 9.1%) of them were in leading roles inside and outside the field of medical imaging. We further investigated the location of these positions and found that the largest sample (n=36, 81.8%) were assigned leadership and administrative roles within their institution, while (n=7, 15.9%) of them had been leading outside their institution. Only one respondent held a leading position inside and outside her institution (Figure 2-E).

**Figure2.**
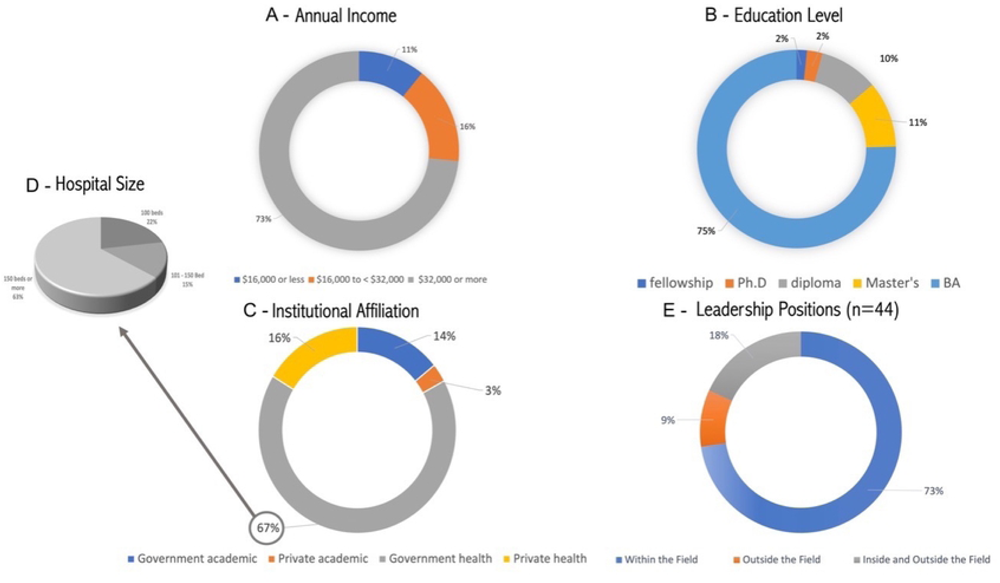

### The Empowerment Factors related to Self-worth

In general, the respondents strongly agreed on self-esteem factors related to leadership roles and administrative positions with an average of 4.29 out of 5.00 and SD=0.622, which indicates the “strongly agree” scale on the study tool. They strongly agree on two factors of self-esteem related roles leadership and administrative which are a) “On the whole, l am satisfied with myself” (85.9%, mean 4.26±0.934), and b) “I feel that I’m a person of worth, at least on an equal plane with others” (95.8%, mean±SD 4.64±0.639). While the rest of the statements had lower strength of agreement compared to the previous two (Table 2). The second axis in the domain of self-efficacy which showed a moderate agreement with an average of 4.04 out of 5.00 that refers to the “I agree” option on the study tool. Breaking down the results we noticed the homogeneity in the agreement on the factors related to this axis, where the average ranged between 3.96 to 4.11. Two factors of self-efficacy are; a) “My coping capability help me calm in a hard situations” ranked first in terms of the agreement of among 80.8% of the respondents with an average of 4.11 out of 5 and SD=0.892, and the second statement b) “ It is easy for me to focus on my objectives and accomplish my goals” ranked second with 73.3% of the total agreement and an average of 3.96 out of 5 SD=0.965.

**Table 2:** Results of Self-Empowerment Dimension.

| Dimension | Statements | Mean | SD |
| --- | --- | --- | --- |
| Self Esteem | Overall, I am satisfied with me | 4.26 | 0.934 |
|  | Overall, I am satisfied with my role in life in general | 4.17 | 0.847 |
|  | I can do things as well as most other people | 4.1 | 0.997 |
|  | I feel that I’m a person of worth, at least on an equal plane with others | 4.64 | 0.639 |
| Self-Efficacy | It is easy for me to stick to my aims and accomplish my goals | 3.96 | 0.965 |
|  | I can remain calm when facing difficulties because I can rely on my coping abilities | 4.11 | 0.892 |

### The Empowerment Factors related to Socio-economical dimension

Most of the study sample agree on most of the economic related factors with an average of 3.78 out of 5.00, which indicates the “I agree” option on the Likert-scale. The highest level of agreement on two statements, a) “I can have/ask for lends and loans” with 79.6% (4.04±0.93), and b) “There is equality in allowance provided from work between men and women (Health and medical field)” with 71.3% (3.87±1). The least agreed statement were; a) “I find equality between men and women in accessing economic opportunities” with 60.5% (3.68±1), and b) “I can benefit various services in the community (such as commercial, investment, and Residential) without restrictions” with 56.2% (3.64±1). Table 3.

**Table 3:** Results of Socioeconomical Dimension.

| Dimension | Statements | Mean | SD |
| --- | --- | --- | --- |
| **Economic Empowerment** | I find equality between men and women in accessing economic opportunities | 3.68 | 1.146 |
|  | I find equality between men and women by giving promotions and awards | 3.69 | 1.227 |
|  | I can have/request for lends and loans | 4.04 | 0.925 |
|  | I can benefit various services in the community (such as commercial, investment, and Residential) without restrictions | 3.64 | 1.013 |
|  | There is an equality in giving an allowance provided from work between men and women (Health and medical field) | 3.87 | 1.083 |

### The Empowerment Factors related to Mobility Freedom

In freedom of **mobility** that can contribute to taking roles in leadership and administrative positions, the general level of agreement was an average of 3.46 out of 5.00, which indicates the “I agree” option. There discrepancy in the agreement on freedom of mobility. Agree on two factors represented in the two phrases; a) “ I can travel inside the Kingdom to participate in conferences and events depending on clear and easy-to-apply submissions” few disagreed while the rest 70.7% agreed on this statement with (3.86±1.065), b) “I can travel outside the Kingdom to participate in conferences and events without restrictions”, had total agreement of only 52.5% (3.47±1.203). However, more than half of the respondent 58.3%(3.06±1.273) did not agree on their freedom of mobility of transfer between jobs in the field of medical imaging which was stated as “I have the freedom of mobility between work fields in radiology with clear policies”, Table 4.

**Table 4:** Results of Mobility Freedom Dimension.

| Dimension | Statements | Mean | SD |
| --- | --- | --- | --- |
| **Mobility Freedom** | I can travel inside the Kingdom to participate in conferences and events depending on clear and easy-to-apply submission | 3.86 | 1.065 |
|  | I can travel outside the Kingdom to participate in conferences and events without restrictions | 3.47 | 1.203 |
|  | I have the freedom of mobility between work fields in radiology with clear policies | 3.06 | 1.273 |

### The Empowerment Factors related to Environmental Factors

The results here are surprising as the female responded to the environmental factors related to empowering women, in general, with neutral response on the Likert-scale with an average of (2.95 out of 5.00). Although many agreed on one factor which is related to the ability to travel freely within the country with 70.5% of total responses (3.49+1.2), they hardly agreed on their freedom to travelling abroad with 52.5% (3.37+1.2). Compared to that, less percentage of female in medical imaging around 44.2% (3.18+1.2) agreed that there is equity in nomination for funding the training courses at work while 29.2% were neutral and 26.7% disagreed with the statement “I find equality between men and women in nominations for courses at work”. It is unfortunate that 50.5-51.5% of these females do not find the support from their work field in keeping gifted, hardworking, and intelligent (2.52+1.2) nor providing a library or resources for updates in the field (2.66+1.3). Table 5 shows the average of responses for the rest of the statements which was the weakest on the population agreement.

**Table 5:** Results of Environmental Dimension.

| Dimension | Statements | Mean | SD |
| --- | --- | --- | --- |
| Environmental factors | I can travel to complete my studies anywhere in Saudi Arabia, according to clear and easy-to-apply conditions and regulations | 3.49 | 1.231 |
|  | I can travel to complete my studies anywhere outside Saudi Arabia, according to clear and easy-to-apply conditions and regulations | 3.37 | 1.199 |
|  | My work provides me with all the needs, tools, equipment, and resources needed to carry out my work effectively | 3.06 | 1.2 |
|  | I get sufficient support to provide initiatives that serve my professional field | 2.94 | 1.124 |
|  | My work supports the professional development through training courses and workshops | 2.79 | 1.278 |
|  | My work supports me to continue postgraduate studies or professional specialization, such as fellowship | 2.74 | 1.31 |
|  | My work provides me with all the needs, tools and resources needed to perform my work effectively | 2.72 | 1.221 |
|  | My employer makes a tangible effort to retain talented employees and encourage them to solve problems in innovative ways | 2.52 | 1.224 |
|  | I find equality between men and women in nominations for courses at work | 3.18 | 1.196 |
|  | My work provides a digital or hard-copy library with research and scientific developments, the latest books and manuals for modern devices | 2.66 | 1.322 |

### The Empowerment Factors related to Institutional factors

The respondents in this section were hesitant to agree or disagree with an average of (3.32 out of 5.00), which is the category that indicates a “neutral” option in the Likert-scale. Responses towards election and nomination for leading positions had higher disagreement 41.3% compared to agreement of 33.3% and neutral 25.4% which pushes the total mean to 3.83+1.3. The next statement that had better level of agreement 66.7% compared to the latter statement is “I feel I do belong to my work field” with an average 3.63+1.2. The rest of the statements’ level of agreement is in Table 6.

**Table 6:** Results of Institutional Dimension.

| Dimension | Statements | Mean | SD |
| --- | --- | --- | --- |
| Institutional factors | I have a real belief in the institutional mission and goals | 3.54 | 1.034 |
|  | I feel I do belong to my work field | 3.63 | 1.146 |
|  | My performance is evaluated impartially | 3.58 | 1.201 |
|  | I have opportunities to learn new skills in my work | 3.16 | 1.21 |
|  | I don't face gender discrimination at the workplace | 3.28 | 1.317 |
|  | I am allowed to elect/nominate for managing and leading positions | 3.83 | 1.264 |
|  | If your boss is absences, the most qualified person is chosen regardless of gender | 3.22 | 1.392 |

### The Empowerment Factors related to job factors

An average of (2.92 out of 5.00) meet the neutral category of the Likert-scale response of females in medical imaging on statements related to job empowering factors that can support women in medical imaging careers. There is significant discrepancy in the agreement among many of the statements. Most of the statements in this dimension is below neutral and towards disagreeing. The agreement level to the rights for breastfeeding hour (47.9%, 3.4+1), rest during working hours (52%, 3.26+1.3), leave for accompanying family member in sickness such as a parent or children (37.5%, 3.06+1.3), sick leaves during pregnancy (36.3%, 3.09+1.2) or monthly period (17%, 2.22+1.2), and the availability for rest area at work had (51.7%, 3.1+1.358). Moreover, childcare services or centres in the institution had more disagreement than the rest of the statements with (63.8%, 2.35+1.3), Table 7.

**Table 7:** Results of Job Empowering Dimension.

| Dimension | Statements | Mean | SD |
| --- | --- | --- | --- |
| Job empowering factors | There is a childcare at my workplace | 2.35 | 1.307 |
|  | There are special places for women to rest at work | 3.1 | 1.358 |
|  | I can get vacations easily during my periods | 2.22 | 1.218 |
|  | I can get vacations during pregnancy | 3.09 | 1.174 |
|  | I can get a sick/medical leave to be with a family member | 3.06 | 1.264 |
|  | I have the right to take advantage of rest hours during official work hours | 3.26 | 1.274 |
|  | I have the right to take advantage of the breastfeeding hour | 3.4 | 1.135 |

### The challenges facing women empowerment

The final dimension was challenges facing women empowerment in medical imaging. Although these challenges were phrased in statements, open question was present in the questionnaire to allow further challenges to be recognised. However, the statements in this dimension had more neutral responses compared to the rest of the scale. Women agree upon the difficulties they face to balance work and life (54.6%, 3.47+1.2) especially the family responsibilities. This may contribute to the long working hours as per more than half of the respondents (52.5%, 3.57+1.2), the societal restriction (42.1%,3.12+1.2), difficulty to care for own children during working hours (41.3%, 3.47+0.95). Less agreement on the statement “Men’s attitude towards women and leadership is negative in the department I work in” (33.4%, 3.1+1.2), Table 8.

**Table 8:** Results of Challenges Towards Women Empowerment.

| Dimension | Statements | Mean | SD |
| --- | --- | --- | --- |
| Challenges | There are social restrictions governing/ controlling my decision-making freedom | 3.12 | 1.198 |
|  | Men's attitude towards women and leadership is negative at the area I work in | 3.1 | 1.162 |
|  | I find it hard to take care of my kids while I'm at work | 3.47 | 0.95 |
|  | My hours are long, and doesn't suit me and my family | 3.57 | 1.187 |
|  | I find it difficult to take care of both my family and work at once | 3.47 | 1.196 |

## Discussion

The study was conducted to investigate women’s status in leadership and administrative positions in the field of medical imaging from both health sector and the academe. Exploring other empowering factors related to these roles and addressing the difficulties facing female empowerment in medical imaging was included.

An important observation emerged from this study is that 67.5% of the sample size were under 30 years of age, which indicate that the number of female graduates has increased recently and that could be attributed to women’s opportunities provided since the announcement of the Saudi 2030 vision [28]. Although previous studies suggest that micro factors such as personal, and familial, factors can influence women’s empowerment, the macro factors such as society, culture, organizations, and governments are more essential to rapidly facilitate empowerment. Self-esteem and self-efficacy are internal factors that can be predictor of women empowerment level [29] which found to be high in our sample (M = 4.29, SD = 0.622) and (M = 4.04, SD = 0.79), respectively. Both factors can be used when designing empowering programs to support women leadership. We consider these results promising for the future empowerment because low self-esteem is a significant obstacle in the work environment as it leads to avoidance of leadership positions and satisfaction by mere participation without the ability to make decisions [25, 30, 31]. Many evidence showed that women faced fewer tenured faculty positions with slower career advancement despite increasing research activity over time, which lead to slower promotion than males [32]. These disparities have persisted for decades, despite increasing in women entering the field of radiology over 40 years, women presence in radiology editorial boards are still lagging and never held a position as editor-in-chief [33].

The economic empowerment is defined as “the transformative process that helps women move from limited power at home to having the skills, resources, and opportunities needed to compete equitably in job markets in order to control and benefit from economic gains” [34]. Our study investigated this domain because women’s access to economic resources and opportunities, such as employment, financial services, real estate, and other productive assets, increases with economic empowerment. We found that the participants were satisfied and agreed on equity between men and women in salaries, allowances, promotions, rewards, and access to investment opportunities, because equal wages are stipulated in the National labor system [35]. Our finding is outstanding and incomparable to the global problem of gender wage inequality where gender pay gaps were reported across all medicine specialties without clear explanation by seniority, career breaks, and/or part-time work [36]. Women inability to negotiate salary results in wage gap in academe [3] which reinforced, along with other studies, that pay gap is one of the international challenges in women empowering [37–40]. Inequality in wages has led many women dropping out of their jobs, which negatively affects women’s empowerment and leadership opportunities. As working in the radiology sector is stressful, some women may find that their financial return when obtaining leadership positions is not worth the efforts [37, 38]. However, our findings in wage equity were not an issue and supports women empowerment in Saudi Arabia.

Besides the economic factors, the mobility freedom for Saudi women in medical imaging was investigated. We found that the participants were satisfied and agreed on the factors of this domain although the women history of mobility freedom had sensational events [41]. In the book “Gender, Class, Nationality, and Women’s Access to Public Places in Riyadh ” Saudi women faced obstacles in many aspects of their lives that hindered their empowerment and independence [42]. Examples of reasons behind the almost complete absence of women in public places mentioned by the author included; a) women driving was prohibited in the past b) the lack of public transportation, c) the financial cost in the case of hiring a driver, d) lack of anti-harassment laws, which is restricting the mobility freedom [42]. Limited access to transportation can increase women’s “hidden” barriers to the participation in the labor market and negatively affect their access to other services such as education or health, and ultimately reduce women’s empowerment [43, 44]. Since the announcement of the 2030 vision, women empowerment is one of the targets and was set to achieve 30% increase women presence in labor market by 2020. Many of these obstacles were resolved and solution is still being facilitated by the government. In 2018, the women were given numerous rights such as allowing them to drive and issue driving licenses besides setting and immediate execution of harassment laws. In 2019, the travel requirement for women with a male guardian was abolished. The *Saudi Human Resources Development Fund* initiated “Wusool” which aims at increase women’s involvement in the job market and support the newbies in overcoming transportation barriers if working for private sector and, thus, ensure job stability. The program provides financial assistance for transportation costs for eligible workers, covering up to 80% of the cost and lasts for 24 months [45]. These changes explain our findings, where participants agreed on the freedom of mobility factors.

We found that although the participants showed a high level of loyalty and passion towards their careers and institutions, no tangible efforts have been made by the workplace in terms of caring and motivating talented and hardworking female employees, therefore the participants showed their dissatisfaction with such element. Studies have confirmed that having talented women in leadership positions benefit the final work outcomes, as well as offer the advantage of different perspectives [46, 47]. Nevertheless, the *Women in the Workplace* 2017 report revealed that although women make up 52% of the US population, they make up only 20% of leadership roles [48, 49]. This reflects how difficult it can be for women to get to the top. During recruitments, institutions can; a) assign diverse committee with prominent females to interview applicants [50], b) showcase the success of female employee/faculty, c) set flexible tone in welcoming environment.

In the context of the institutional empowerment, participants agreed that workplace lack of nurseries and childcare, prohibit leave during menstrual cycle. One practical factor that assists women in fully participating in the workplace is the implementation of institutional policies that promote exclusion of the gendered experience. Inclusive policies which help foster an enabling environment are those that provide support for working female parents and continually evaluate workplace flexibility. Qurrah Subsidy Program is another Saudi initiative that was launched through the Child Welfare Authority intended to provide support to working mothers by ensuring that their children are well-cared for while they pursue employment opportunities in private sector [51]. This explains the increase in the number of fresh women in the field of medical imaging by our findings.

As for the difficulties in the field of medical imaging, long working hours, lack of development, and the inability to attend conferences and workshops are among the most common barriers that were repeated by the participants. Professional conferences are an important source of training, networking, and learning opportunities in every field [52]. To limit the negative impact of these barriers, we propose part-time job opportunities and motivate stand-out workers at a rate of one workshop per month for the most accomplished employee. We call for the establishment of programs that promote the development of women’s skills in management and leadership, as their effectiveness has been evident [53]. Having an administrative role in the field of medical imaging reflects empowering status and indicates effective women leadership and influence in the field. This enables role modelling function for future generations. Unexpectedly, our study revealed that majority of our sample (81.7%) have never held a leadership or administrative position. Which is supported by underrepresented US academic women in the field of radiology despite having comparable levels of experience to men [54, 55]. Workplace is expected to provide training and development programs for employees/faculty on topics such as unconscious prejudice, negotiation skills, and equity. The low presence of women in leading positions in our study may be attributed to two reasons; 49.4% of the participants were recent graduates, and 60.4 % of our sample have less than 5 years of work experience. This supported by previous literature where higher number of women fill low rank positions compared to advanced ranks [56, 57]. Women are 18% less likely to get promoted than their male peers [46]. Without supportive policies, talented employees will fall behind or not being able to fully reach maximum potential in their careers [37, 46].

Another attribute to underrepresentation in leading positions and advanced ranks, is the lack of retention of women particularly in academia [58]. Lack of role models and poor mentoring can hinder female to accept leading/administrative positions [59]. Active support in the workplace through mentoring talented women to achieve their full potential is recommended to avoid the “leaky pipeline” which is pronounced in medical fields [60]. Further evident reason is that women miss such opportunities due to familial obligations and societal expectations following parenting and domestic responsibilities [61]. We recommend implementing family-friendly policies and promoting a work– life integration culture at institutional, departmental, and individual levels tailored to acknowledge gender-specific difficulties [62]. These can be achieved through f1exible working hours and support for returners after maternity breaks or leaves (Table 9). Workforce policies should include provisions for breastfeeding or lactation breaks for new mothers. The Saudi working woman has the right for a conditional paid maternity leave up to three years after birth. If returning to work before that, she is entitled to a period, or rest periods, for the purpose of breastfeeding her newborn, with pay, not exceeding a total of an hour per day for 24 months postpartum [63].

**Table 9:**
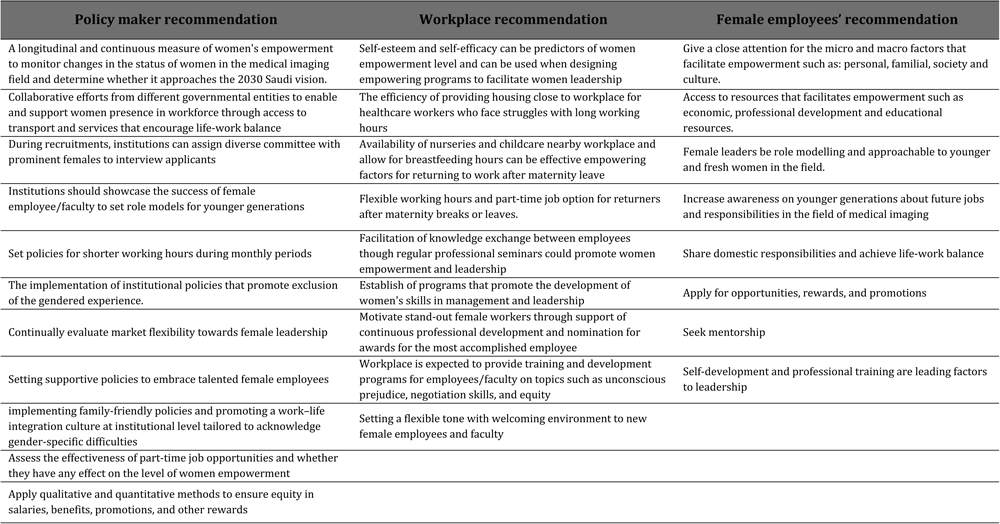
Summary of recommendations to empower women and promote equity.

A limitation of our study is that many participants have recently graduated and lack work experience, and this is attributed to the tremendous changes that Saudi Arabia is going through to achieve Vision 2030 through offering academic programs for female that was previously inclusive to male and enforcing more job opportunities for women. These are not surprising compared to global evidence of increases drop-off of academic females at the associate and full professor levels [59]. On top of all, the pandemic has affected women more than men, and it impede the promotion of female academic radiologists as it has adversely affected academics’ productivity and disproportionately increased women’s household and childcare duties [64]. Further studies are required to measure the extent, efficacy, and quality of women empowerment in leadership and administrative positions.

The largest increase in the representation of women in leadership positions in tight cultures should come through egalitarian behaviors. There should be a strong dedication by decision makers to significantly improve women status in empowering positions and avoid the implicit biases in women leadership prototypes. Future research is mandated in the effectiveness of women leadership in this field and specifically in the Middle East and North Africa (MENA) and developing countries. In addition, researchers ought to explore the gender diversity in fields where women are underrepresented to come up with a more proactive strategy for laying the foundation for gender parity in the burgeoning industries of the future. We need to establish promoting women for leadership positions in the field of medical imaging academe and industry to facilitate the placement of highly qualified women into leadership positions.

## Conclusion

The current status of women in the field of medical imaging and the challenges that could lead to the glass ceiling for women empowerment in both sectors is presented here. Our study found that women do not lack self-esteem or efficacy and government supports their presence and empower them. The culture places some constraints on women emancipation, however, the real hurdle lays upon the recruiting institutions to facilitate the emergence of women as leaders. The glass ceiling effect is an existing issue bolstering female leadership in fields dominated by men through accelerating gender egalitarianism must be a core part of the public and private agenda. The Saudi Arabian government has demonstrated exemplary leadership of proactive initiatives that effectively empowered women in the workforce.

## Data Availability

All relevant data are within the manuscript and its Supporting Information files.

## Acknowledgments

We extend our deepest gratitude to the remarkable women who generously participated in this research study. Your voices have added depth to our findings, and we hope that the insights gained from this study will contribute to the betterment of women empowerment all around the world.

## References

1. Alghofaily L. Women leadership in higher education in Saudi Arabia. International Journal of Social Sciences. 2019;8(2):14–32.

2. UNESCO. Just 30% of the world’s researchers are women. What’s the situation in your country? : UNESCO; 2021 [cited 2023 02/01/2023]. Available from: https://shar.es/aflDVp.

3. Shannon G, Minckas N, Tan D, Haghparast-Bidgoli H, Batura N, Mannell J. Feminisation of the health workforce and wage conditions of health professions: an exploratory analysis. Hum Resour Health. 2019;17(1):72. Epub 2019/10/19. doi: 10.1186/s12960-019-0406-0. PubMed PMID: 31623619; PubMed Central PMCID: PMCPMC6796343.

4. Schor NF. The Decanal Divide: Women in Decanal Roles at U.S. Medical Schools. Academic Medicine. 2018;93(2).

5. Al shammary SE, Zrieq R, Ibrahem UM, Altamimi AB, Diab HM. On the Celebration of the International Day of Women and Girls in Science: Assessment of the Factors Mediating Women’s Empowerment in Scientific Research in Saudi Arabia. Sustainability. 2021;13(4):2385. PubMed PMID: 10.3390/su13042385.

6. Hay K, McDougal L, Percival V, Henry S, Klugman J, Wurie H, et al. Disrupting gender norms in health systems: making the case for change. The Lancet. 2019;393(10190):2535–49. doi: 10.1016/S0140-6736(19)30648-8.

7. Ahmed W. Women Empowerment in Saudi Arabia: An Analysis from Education Policy Perspective. The Middle East International Journal for Social Sciences (MEIJSS). 2020;2(3):93–8.

8. Rajeh M, Nicolau B, Qutob A, Pluye P, Esfandiari S. A Survey of the Barriers Affecting the Career Practice and Promotions of Female Dentists in Saudi Arabia. JDR Clinical & Translational Research. 2019;4(2):187–95. doi: 10.1177/2380084418815458.

9. Rajeh M, Nicolau B, Pluye P, Qutob A, Esfandiari S. Are There Barriers for Professional Development of Women Dentists? A Qualitative Study in Saudi Arabia. JDR Clinical & Translational Research. 2017;2(2):119–31. doi: 10.1177/2380084416685083.

10. Gupta D, Garg J. Sexual harassment at workplace. International Journal of Legal Science and Innovation; 2020.

11. Gaines J. Women in male-dominated careers. 2017.

12. Heiser S. The Majority of U.S. Medical Students Are Women, New Data Show. 2019.

13. Boakye AO, Dei Mensah R, Bartrop-Sackey M, Muah P. Juggling between work, studies and motherhood: The role of social support systems for the attainment of work–life balance. SA Journal of Human Resource Management. 2021;19:10.

14. Mason K, Smith H. Women’s Empowerment and Social Context: Results from Five Asian Countries. 2012.

15. Miedema SS, Haardörfer R, Girard AW, Yount KM. Women’s empowerment in East Africa: Development of a cross-country comparable measure. World Development. 2018;110:453–64. doi: 10.1016/j.worlddev.2018.05.031.

16. Kemper KJ, Schwartz A, Consortium PRB-RS. Bullying, discrimination, sexual harassment, and physical violence: common and associated with burnout in pediatric residents. Academic pediatrics. 2020;20(7):991–7.

17. SteelFisher GK, Findling MG, Bleich SN, Casey LS, Blendon RJ, Benson JM, et al. Gender discrimination in the United States: Experiences of women. Health Services Research. 2019;54(S2):1442–53. doi: 10.1111/1475-6773.13217.

18. Sathyasri B, Vidhya UJ, Sree GJ, Pratheeba T, Ragapriya K. Design and implementation of women safety system based on Iot technology. International Journal of Recent Technology and Engineering (IJRTE). 2019;7(6S3).

19. Mason KO, Smith HL. Women’s empowerment and social context: Results from five Asian countries. Gender and Development Group, World Bank, Washington, DC. 2003;53(9).

20. Akter S, Rutsaert P, Luis J, Htwe NM, San SS, Raharjo B, et al. Women’s empowerment and gender equity in agriculture: A different perspective from Southeast Asia. Food Policy. 2017;69:270–9. doi: 10.1016/j.foodpol.2017.05.003.

21. World Economic Forum. Global Gender Gap Reports. Report. Switzerland: 2022.

22. Agarwal B. Gender equality, food security and the sustainable development goals. Current Opinion in Environmental Sustainability. 2018;34:26–32. doi: 10.1016/j.cosust.2018.07.002.

23. Sen G, George A, Ostlin P, Ramos S. Unequal, Unfair, Ineffective and Inefficient Gender Inequity in Health: Why it exists and how we can change it. 2007.

24. Al-Amoudi SM. Health empowerment and health rights in Saudi Arabia. Saudi Med J. 2017;38(8):785–7. doi: 10.15537/smj.2017.8.19832. PubMed PMID: 28762428; PubMed Central PMCID: PMCPMC5556292.

25. Al-Qahtani AM, Ibrahim HA, Elgzar WT, El Sayed HAE, Abdelghaffar T, Moussa RI, et al. Perceived and real barriers to workplace empowerment among women at Saudi universities: A cross-sectional study. Afr J Reprod Health. 2021b;25(s1):26–35. Epub 2021/06/03. doi: 10.29063/ajrh2021/v25i1s.3. PubMed PMID: 34077142.

26. Unified National Platform. Women Empowerment: GOV.sa; 2021 [updated 07/07/2021; cited 2023 3 Feb 2023]. Available from: https://www.my.gov.sa/wps/portal/snp/careaboutyou/womenempowering/!ut/p/z0/04_Sj9CPykssy0xPLMnMz0vMAfIjo8zijQx93d0NDYz8LYIMLA0CQ4xCTZwN_Ay8TIz0g1Pz9AuyHRUBwQYLNQ!!/#:∼:text=The%20Kingdom%20advanced%20in%20a,and%20receiving%2080%2F100%20points.

27. Shannon G, Jansen M, Williams K, Cáceres C, Motta A, Odhiambo A, et al. Gender equality in science, medicine, and global health: where are we at and why does it matter? The Lancet. 2019;393(10171):560–9. doi: 10.1016/S0140-6736(18)33135-0.

28. Nuruzzaman M. Saudi Arabia’s ’Vision 2030’: Will It Save Or Sink the Middle East? : E-International Relations; 2018. Available from: https://www.e-ir.info/2018/07/10/saudi-arabias-vision-2030-will-it-save-or-sink-the-middle-east/.

29. Al-Qahtani AM, Ibrahim HA, Elgzar WT, El Sayed HA, Essa RM, Abdelghaffar TA. The role of self-esteem and self-efficacy in women empowerment in the Kingdom of Saudi Arabia: A cross-sectional study. Afr J Reprod Health. 2021a;25(s1):69–78. Epub 2021/06/03. doi: 10.29063/ajrh2021/v25i1s.7. PubMed PMID: 34077146.

30. Adjei SB. Assessing Women Empowerment in Africa: A Critical Review of the Challenges of the Gender Empowerment Measure of the UNDP. Psychology and Developing Societies. 2015;27(1):58–80. doi: 10.1177/0971333614564740.

31. Offermann LR, Beil C. Achievement Styles of Women Leaders and Their Peers: Toward an Understanding of Women and Leadership. Psychology of Women Quarterly. 1992;16(1):37–56. doi: 10.1111/j.1471-6402.1992.tb00238.x.

32. Jalilianhasanpour R, Chen H, Caffo B, Johnson P, Beheshtian E, Yousem DM. Are Women Disadvantaged in Academic Radiology? Academic Radiology. 2020;27(12):1760–6. doi: 10.1016/j.acra.2020.09.019.

33. Piper CL, Scheel JR, Lee CI, Forman HP. Representation of Women on Radiology Journal Editorial Boards: A 40-Year Analysis. Academic Radiology. 2018;25(12):1640–5. doi: 10.1016/j.acra.2018.03.031.

34. WEE. Women’s Economic Empowerment: Bill & Melinda Gates Foundation; 2017 [November 4, 2022]. Available from: https://www.gatesfoundation.org/equal-is-greater/our-approach/.

35. Alkhudair D. Saudi Arabia looks to close gender pay gap: Arab News; 2020 [cited 2022 27/11/2022]. Available from: https://arab.news/vbqdx.

36. Connolly S, Holdcroft A, editors. The pay gap for women in medicine and academic medicine2009.

37. Fichera G, Busch IM, Rimondini M, Motta R, Giraudo C. Is Empowerment of Female Radiologists Still Needed? Findings of a Systematic Review. Int J Environ Res Public Health. 2021;18(4). Epub 2021/02/11. doi: 10.3390/ijerph18041542. PubMed PMID: 33562881; PubMed Central PMCID: PMCPMC7915271.

38. Surawicz CM. Women in Leadership: Why So Few and What to Do About It. J Am Coll Radiol. 2016;13(12 Pt A):1433-7. Epub 2017/03/28. doi: 10.1016/j.jacr.2016.08.026. PubMed PMID: 28341310.

39. Weigel KS, Kubik-Huch RA, Gebhard C. Women in radiology: why is the pipeline still leaking and how can we plug it? Acta Radiologica. 2019;61(6):743–8. doi: 10.1177/0284185119881723.

40. Witteman HO, Hendricks M, Straus S, Tannenbaum C. Are gender gaps due to evaluations of the applicant or the science? A natural experiment at a national funding agency. Lancet. 2019;393(10171):531–40. Epub 2019/02/12. doi: 10.1016/s0140-6736(18)32611-4. PubMed PMID: 30739688.

41. Geel A. Separate or together? Women-only public spaces and participation of Saudi women in the public domain in Saudi Arabia. Contemporary Islam. 2016;10. doi: 10.1007/s11562-015-0350-2.

42. Le Renard A. Genre, classe, nationalité et accès des femmes aux espaces publics à Riyad. Sociétés contemporaines. 2011;84(4):151–72. doi: 10.3917/soco.084.0151.

43. Dulhunty A. Disciplinary mobility and women’s empowerment: a complicated connection. Mobilities. 2022:1–15. doi: 10.1080/17450101.2022.2088298.

44. Sharma-Brymer V, Sharma SNV. Women and Transport: A Comparative Analysis of Issues and Actions. In: Leal Filho W, Marisa Azul A, Brandli L, Lange Salvia A, Wall T, editors. Gender Equality. Cham: Springer International Publishing; 2021. p. 1129-39.

45. Wusool. “Wosool” program for fund transporting working women: Saudi Human Resources Development Fund; 2020 [cited 2023 March, 3]. Available from: https://wusool.sa/service.html.

46. Odei BC, Seldon C, Fernandez M, Rooney MK, Bae J, Acheampong J, et al. Representation of Women in the Leadership Structure of the US Health Care System. JAMA Network Open. 2021;4(11):e2136358-e. doi: 10.1001/jamanetworkopen.2021.36358.

47. Ruderman MN, Ohlott PJ, Panzer K, King SN. Benefits of Multiple Roles for Managerial Women. Academy of Management Journal. 2002;45(2):369–86. doi: 10.5465/3069352.

48. Hiraoka AA, Bohrer A, Gina C, Deb C, Marianne C, Christianne C, et al. Women in the Workplace 2022: LeanIn.Org and McKinsey & Company 2022. Available from: https://womenintheworkplace.com.

49. Krivkovich A, Robinson K, Starikova I, Valentin R, Yee L. Women in the workplace 2017. 2017.

50. Sepulveda KA, Paladin AM, Rawson JV. Gender Diversity in Academic Radiology Departments: Barriers and Best Practices to Optimizing Inclusion and Developing Women Leaders. Academic Radiology. 2018;25(5):556–60. doi: 10.1016/j.acra.2017.08.018.

51. Qurrah SP. “Qurrah” Subsidy Program a national initiative of the Child Welfare Authority: Human Resources Development Corporation 2017. Available from: https://qurrah.sa/aboutsubsidy.

52. Ngamsom B, Beck J. A Pilot Study of Motivations, Inhibitors, and Facilitators of Association Members in Attending International Conferences. Journal of Convention & Exhibition Management. 2000;2(2-3):97–111. doi: 10.1300/J143v02n02_09.

53. Spalluto LB, Spottswood SE, Deitte LA, Chern A, Dewey CM. A Leadership Intervention to Further the Training of Female Faculty (LIFT-OFF) in Radiology. Acad Radiol. 2017;24(6):709–16. Epub 2017/05/21. doi: 10.1016/j.acra.2016.12.025. PubMed PMID: 28526513.

54. Goswami AK, Kokabi N, Khaja MS, Saad WE, Khaja A, Vashi AP, et al. Academic Radiology in the United States: Defining Gender Disparities in Faculty Leadership and Academic Rank. Academic Radiology. 2022;29(5):714–25. doi: 10.1016/j.acra.2021.05.016.

55. Fichera G, Busch IM, Rimondini M, Motta R, Giraudo C. Is Empowerment of Female Radiologists Still Needed? Findings of a Systematic Review. International Journal of Environmental Research and Public Health. 2021;18(4):1542. PubMed PMID: 10.3390/ijerph18041542.

56. Qamar SR, Khurshid K, Jalal S, McInnes MDF, Probyn L, Finlay K, et al. Gender Disparity Among Leaders of Canadian Academic Radiology Departments. American Journal of Roentgenology. 2019;214(1):3–9. doi: 10.2214/AJR.18.20992.

57. Hamidizadeh R, Jalal S, Pindiprolu B, Tiwana MH, Macura KJ, Qamar SR, et al. Influences for Gender Disparity in the Radiology Societies in North America. American Journal of Roentgenology. 2018;211(4):831–8. doi: 10.2214/AJR.18.19741.

58. Carr PL, Gunn CM, Kaplan SA, Raj A, Freund KM. Inadequate Progress for Women in Academic Medicine: Findings from the National Faculty Study. Journal of Women’s Health. 2015;24(3):190–9. doi: 10.1089/jwh.2014.4848.

59. Surawicz C. Women in Leadership: Why So Few and What to Do About It. Journal of the American College of Radiology. 2016;13(12, Part A):1433-7. doi: 10.1016/j.jacr.2016.08.026.

60. Weigel KS, Kubik-Huch RA, Gebhard C. Women in radiology: why is the pipeline still leaking and how can we plug it? Acta Radiologica. 2020;61(6):743–8. doi: 10.1177/0284185119881723. PubMed PMID: 31648538.

61. Jolly S, Griffith KA, DeCastro R, Stewart A, Ubel P, Jagsi R. Gender differences in time spent on parenting and domestic responsibilities by high-achieving young physician-researchers. Annals of internal medicine. 2014;160(5):344–53.

62. Agrawal P, Madsen TE, Lall M, Zeidan A. Gender Disparities in Academic Emergency Medicine: Strategies for the Recruitment, Retention, and Promotion of Women. AEM Education and Training. 2020;4(S1):S67–S74. doi: 10.1002/aet2.10414.

63. HRSD. Medical care for pregnancy and childbirth, and the rest period after maternity leave Saudi Arabia: Human Resources and social development; 2023. Available from: http://laboreducation.hrsd.gov.sa/ar/labor-education/287.

64. Tso HH, Parikh JR. Mitigating delayed academic promotion of female radiologists due to the COVID pandemic. Clinical Imaging. 2021;76:195–8. doi: 10.1016/j.clinimag.2021.04.010.

